# Estimation of Quasi-Continuous Blood Pressure based on Harmonic Phase-Shifts in Signals using Non-Invasive Photoplethysmographic Measurements

**DOI:** 10.1101/2019.12.19.19015321

**Authors:** Fabian Kern, Stefan Bernhard

## Abstract

According to the guidelines of the European Society of Hypertension International Protocol revision 2010, the requirements for long time blood pressure measurement (BPM) are: simple handling, robustness against movements, accuracy of better than *±* 5 mmHg, and, above all, that the patient’s motion should not be restricted during measurement. These requirements are in particular important for a reliable interpretation of the blood pressure (BP) of hypertensive patients, because such a diagnosis will usually be confirmed by long-term measurement. Moreover, to be able to correlate the patient’s BP with his normal daily activity, non-obstructive non-invasive methods are desired to reduce the patient’s load.

The main concern of this paper is to present a novel method for estimating non-invasive continuous blood pressure (CBP) from a single photoplethysmography (PPG) signal. In contrast to the pulse transit time (PTT) method, our approach is based on the assumption that the phase-velocities of the fundamental and higher harmonics depend on the (non-linear) elastic properties of the arteries. Consequently, phase velocity varies as a function of a vessel’s instantaneous dilation and can be effectively utilised for CBP estimation.

In addition to its numerous advantages for a simplified measurement setup, we could show that the method achieves a high degree of correlation for a reliable BP estimation from PPG data. Comparison with state-of-the-art PTT methods was carried out using a dataset from the PhysioBank Database comprising a reference invasive blood pressure (IBP) signal measured at the radial artery, a PPG signal measured at the fingertip and a standard ECG signal.

The correlation values obtained from the long-time estimation of the systolic blood pressure (SBP) were as high as r = 0.8945, while the value for the diastolic blood pressure (DBP) was found to be r = 0.9082 and the correlation of the mean blood pressure (MBP) was r = 0.9322. These results were achieved by analysing the dataset in a beat-to-beat manner and regarding several post-processing procedures like coherent averaging (CA) and zero padding with quasi-continuous frequency domain estimation and artificially refined frequency resolution.

## 1. Introduction

Blood pressure as a vital sign is regarded as the most important parameter of the cardiovascular system (CVS), and it is commonly used as an indicator of general health, especially for people of advanced age, who have a higher risk of developing heart diseases. Furthermore, high BP is known to increase the risk of cardiovascular disease which is the main cause of death worldwide [1].

To ensure a reliable diagnosis of hypertension, CBP is performed over a period of 24 hours and under normal conditions. Further, non-invasive non-obstructive methods are preferred that reduce the patient’s discomfort as much as possible. An overview about the different BPM methods and their advantages and disadvantages is given in [2–4]. Cuff-based methods usually require at least two minutes of spare time between subsequent measurements to reduce measurement errors caused by the external compression of the tissue. While their results are not continuous in time, they put a high burden to the patient. Even if the number of inflations during a 24h measurement are perceived to be quite frequent, the resulting BP time series has a very low time resolution. Continuous beat-to-beat measurements of BP obtained by the computation of the PTT from electrocardiography (ECG) and peripheral PPG signals however, are disputable for several reasons: (i) the dependency on the location and the distance of measurement points, (ii) the method used to find and interpret minima, maxima and saddle points within the PPG and ECG signals [5, 6], (iii) the appearance of undetermined fluctuations in the time between the pressure wave and the ECG R-peak during blood ejection of the left ventricle [7] and (iv) the influences of auto-regulation on arterial stiffness that produce undetermined drifts of the PTT over time [8]. Nonetheless, the PTT method is subject of several publications that aim at a development of wearable continuous BPM devices [5, 9–13]. As PTT-based methods compute the BP by employing its correlation with the actual pulse wave velocity (PWV), the particular drawback of this approach is that PWV depends on the elasticity of the tested artery section, which in turn is considerably affected by a variety of physiological mechanisms of BP regulation.

The general relationship between BP and PTT was subject of a series of investigations. In [6, 14–22] different magnitudes for the computation of the PTT are based on the calculation of the time difference between the ECG R-peak and some reference within the PPG signal and are methodically described and validated by calculating a correlation to the IBP. In addition to the methodological aspects of the BP computation using the PTT, [23] also discusses the impact of influencing factors, like changes in individual arterial stiffness. It was found that the method could sufficiently satisfy the requirements of BP monitoring during sleep, according to low regulation activity of the arteries. The method was tested on ten subjects with errors less than 8% in relation to the reference mean arterial pressure. Furthermore, the correlation of the PTT and the SBP/DBP was investigated for different patient groups of 10 to 64 subjects with all ages and health conditions [5, 24–27]. In the framework of these investigations, the correlation coefficient *r* of the PTT and the SBP was found to be in the range 0.73 ≤ |*r*| ≤ 0.95.

However, all currently available methods require the analysis of at least two signals, the ECG and the PPG, to compute the PTT. Moreover, since the resulting PTT strongly depends on some choice analysis, it remains unclear how the optimal measurement location and the reference points within the time series are determined.

In this paper, we present a new method to determine the continuous BP from a single standard PPG signal measured at the fingertip and its application to a dataset taken from [28] that comprises PPG, ECG and IBP signals, all measured in-sync and with a sample rate of *f*_*s*_ = 125 Hz. In contrast to previously proposed methods, we analyse the PPG signal in the frequency domain and compute the harmonic phase shift between the fundamental and the first harmonic frequency. This phase-shift is caused by the non-linear compliance of the arteries, which has the effect that a wave with a larger amplitude propagates at a higher phase velocity than a wave with a smaller amplitude. With phase-shift related to compliance and compliance related to pressure, the correlation of phase-shift and BP can be calculated and exploited. Finally, the correlation results of the proposed method are compared to the standard PTT procedure.

## 2. Methods

Non-linear arterial compliance is known to be the physiological basis to predict the BP from changes in PTT resulting from a change of PWV along some arterial section. The proposed method, however, analyses phase-shifts in the frequency domain caused by a similar effect. With the fundamental frequency determined by the heart rate (HR), the increase of PWV at higher pressures also goes along with relative phase-shifts between the frequencies in the spectrum of the waveform found in the PPG signal.

Physiological regulation processes neglected, any change in BP induces a change in artery stiffness, expressed as incremental elastic modulus *E* (*E* (*p*)). Equation 1, known as Moens-Korteweg Equation, describes the relationship between the PWV and the geometrical and elastic parameters, where *E* (*p*) is the circumferential elongation of the artery according to changes in transmural pressure *p. h* and *d* denote the vessel’s wall thickness and diameter respectively and *ρ* the fluid density [24, 29–35].

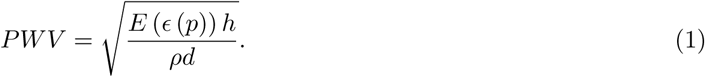

Arterial wall stiffness which is expressed by the incremental elastic modulus *E* (*ϵ*(*p*)) is further known to increase exponentially with mean distending pressure [32]. With PWV depending on the BP, the phase velocities of the components in the pulse wave spectrum are further affected by instantaneous pressure.

The Fourier spectrum of the (continuous) PPG signal particularly includes the fundamental and its first harmonic, with the amplitude of the fundamental being significantly larger than the amplitude of the harmonic. Consequently, any change in BP that alters the elastic modulus will affect the fundamental frequency more than the harmonic, and thus cause a mutual phase-shift that can be quantified by spectrum analysis and correlated with the BP. Evidently this method can be conducted with any other pair of frequencies contained in the spectrum that has significantly differing characteristics. For an alternative approach, Soliton theory would offer itself to describe this relationship in terms of the propagation speed of weakly non-linear waves in a tube in time domain [36].

In the following, we describe in more detail how the signal is divided into subsequent heartbeats and how the fundamental frequency and the first harmonic are determined by use of the discrete Fourier transform (DFT) in order to compute the mutual phase-shift between these two frequencies. Second, we introduce and evaluate several post-processing methods that optimise the results in terms of time resolution and correlation.

### 2.1. Data Basis

The proposed method is applied using a dataset from the “UCI Machine Learning Repository” of the University of California, Irvine [37], which is part of the “PhysioNet” data base [38]. The dataset consists of three vital signals (PPG, ECG and IBP), all sampled at a rate of 125 Hz over a total time of 8m and 57s. In order to correct the time shift caused by the measurement setup, the PPG signal is shifted by 296 ms. During the measurement the SBP varied between 106 mmHg and 180 mmHg, while the DBP and HR ranged from 54 mmHg to 75 mmHg and 74 bpm to 107 bpm respectively. The MBP is calculated as the arithmetic mean of systolic and diastolic blood pressure. In total, the signal extends over *N*_*p*_ = 843 heartbeats. The dataset therefore covers reasonable conditions generally observed in hypertensive patients.

### 2.2. Determination of Beat-specific Parameters for Further Analysis

Due to the quasi-periodic nature of the vital signals, the method is constructed to work with single periods in a beat-to-beat manner only. It therefore requires the segmentation of the signal into a sequence of pulse waves that allows for a further phase-shift analysis for every single heartbeat. The ECG signal is particularly suitable for this, because the R-peak is supposed to indicate the starting point of the blood ejection into the aorta reliably. Furthermore, R-peak detection is quite simple and precise from a numerical point of view [39, 40]. The method we used for segmentation simply determined the maxima above a certain level and was sufficient with respect to the pre-processing, smoothness and characteristics of the regarded data. In contrast to available PTT methods, the R-peak merely serves us as a time marker for separating consecutive heartbeats within the PPG signal. The R-peak serves as an exact specification of the period length, the actual dissection of the periods is done 37 samples after the R-peak as shown in Figure 1. In order to compare the actual results with the PTT method, the PTT is calculated for every beat. In this case, the PTT are calculated from the R-peak to the minimum of the PPG signal, as well as the inflexion point and to the maximum of the PPG and then correlated with the BP.

**Figure 1:**
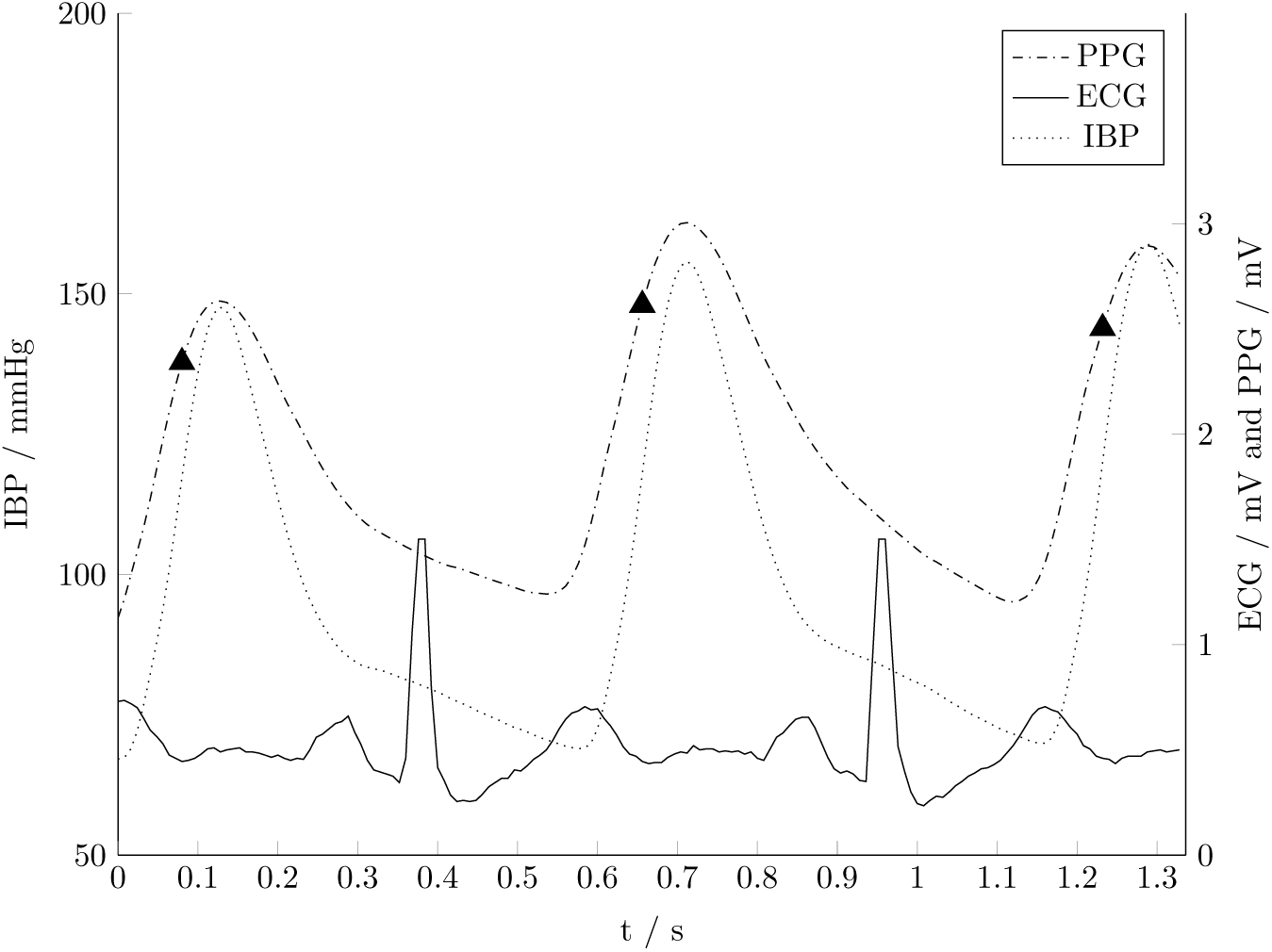
Use of the ECG signal with a constant delay of 37 samples to split the IBP (a. radialis) and PPG (fingertip) signals for further beat-to-beat analysis. For simple and accurate period determination, consecutive R-peaks within the ECG signal were used as detection criterion. The marker ▴ indicates the splits.

In order to compare the results of the novel method with the state-of-the-art PTT method, the PTT is calculated and correlated with the BP. Therefore three different PTT are calculated beginning from the R-peak to the i) minimum, ii) inflexion point and iii) maximum of the PPG signal. The correlation coefficients of the three PTT methods with the SBP, MBP and DBP are shown in table 1.

**Table 1:** Overview of the linear correlation coefficients for the relation of the IHPS and the SBP, MBP and DBP achieved with the different post processing methods.

| Method | Correlation<br>coefficient $r_{max}$<br>with SBP | Correlation<br>coefficient $r_{max}$<br>with MBP | Correlation<br>coefficient $r_{max}$<br>with DBP |
| --- | --- | --- | --- |
| $PTT_{min}$ | -0.3805 (1 period) | -0.4177 (1 period) | -0.4341 (1 period) |
| $PTT_{TP}$ | -0.5558 (1 period) | -0.5931 (1 period) | -0.6119 (1 period) |
| $PTT_{max}$ | -0.5984 (1 period) | -0.6296 (1 period) | -0.6471 (1 period) |
| Single-period | 0.8600 (1 period) | 0.9002 (1 period) | 0.8391 (1 period) |
| Multi-period | 0.8837 (5 periods) | 0.9146 (5 periods) | 0.8742 (8 periods) |
| Coherent averaging without overlapping | 0.8945 (15 periods) | 0.9322 (15 periods) | 0.9082 (11 periods) |
| Zero padding with quasi continuous spectrum | 0.5778 (1 period) | 0.5828 (1 period) | 0.4849 (1 period) |

Furthermore, the following instantaneous parameters were calculated and analysed:

1. Pulse transit time (PTT)
2. Instantaneous heart rate (IHR)
3. Instantaneous systolic blood pressure (ISBP)
4. Instantaneous diastolic blood pressure (IDBP)
5. Instantaneous phase-shifts between the fundamental and the first harmonic frequency of the PPG signal (IHPS)

### 2.3. Correlation of the IHPS to BP

Suspected linear interdependence between two signals can be quantified by calculating the empirical linear correlation coefficient *r*. Given by

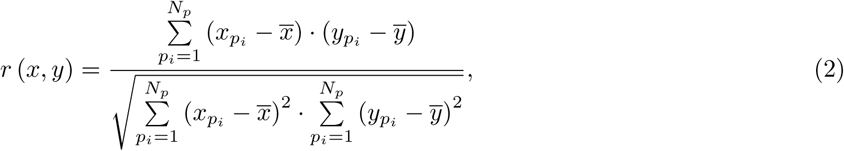

this correlation coefficient is a measure for the similarity of two signals *x* and *y*, where 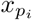 and 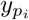 are the instantaneous values, 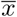 and 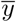 are the mean values, *p*_*i*_ is the period index and *N*_*p*_ is the total number of periods.

According to equation 2, the correlation coefficient of two signals is always in the range −1 ≤ *r* ≤ 1, whereby a value |*r*| *>* 0.8 is considered to indicate a strong correlation between two variables [41].

Apparently, the beat-oriented analysis of the signal has the disadvantage that the individual heartbeats differ in time length. Therefore, the frequency resolution Δ*f*_*i*_ of the DFT can fluctuate from beat to beat and influence the detection of the harmonics. The length variation can be seen in Figure 3 showing the instantaneous heart rate (IHR) in beats per minute and in context with the IHPS and the SBP. It calculates as

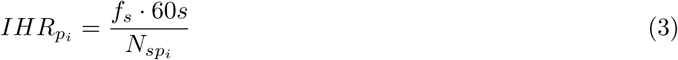

from the sampling frequency *f*_*s*_ and the number 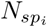 of sampled values attributed to the *i*^th^ period *p*_*i*_. With *r* (*IHR, DBP*) = 0.8774, *r* (*IHR, MBP*) = 0.9305 and *r* (*IHR, SBP*) = 0.8936, the empirical correlations between these variables were reasonably strong, which also becomes evident by the similarities of the graphs in Figure 3.

**Figure 2:**
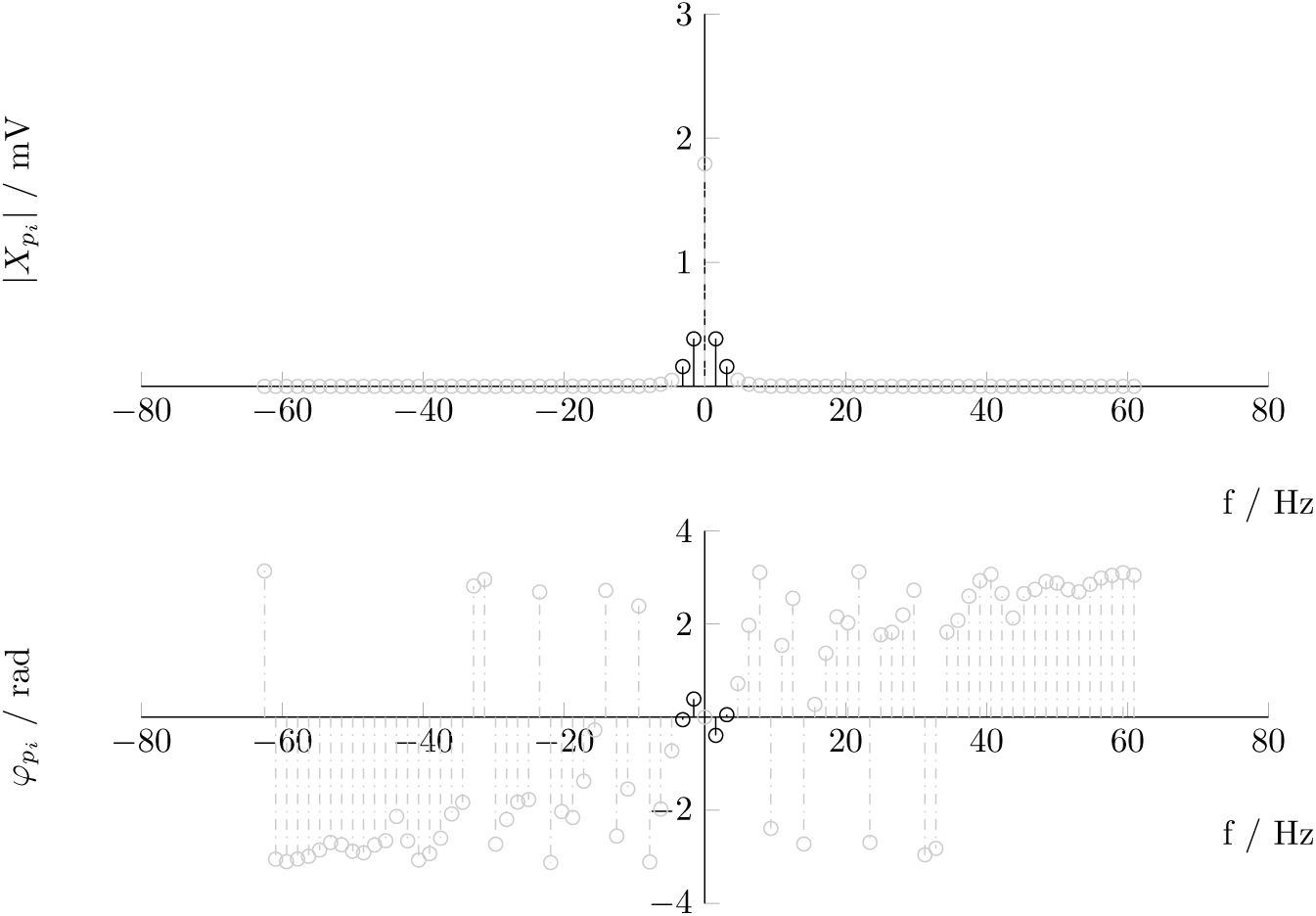
Amplitudes 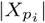 (upper diagram) and phases 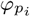 (lower diagram) of a single pulse wave within the PPG signal plotted as a double-sided frequency spectrum. The amplitudes and phases of the fundamental 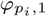 and the first harmonic 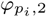 of the signal are highlighted by solid lines.

**Figure 3:**
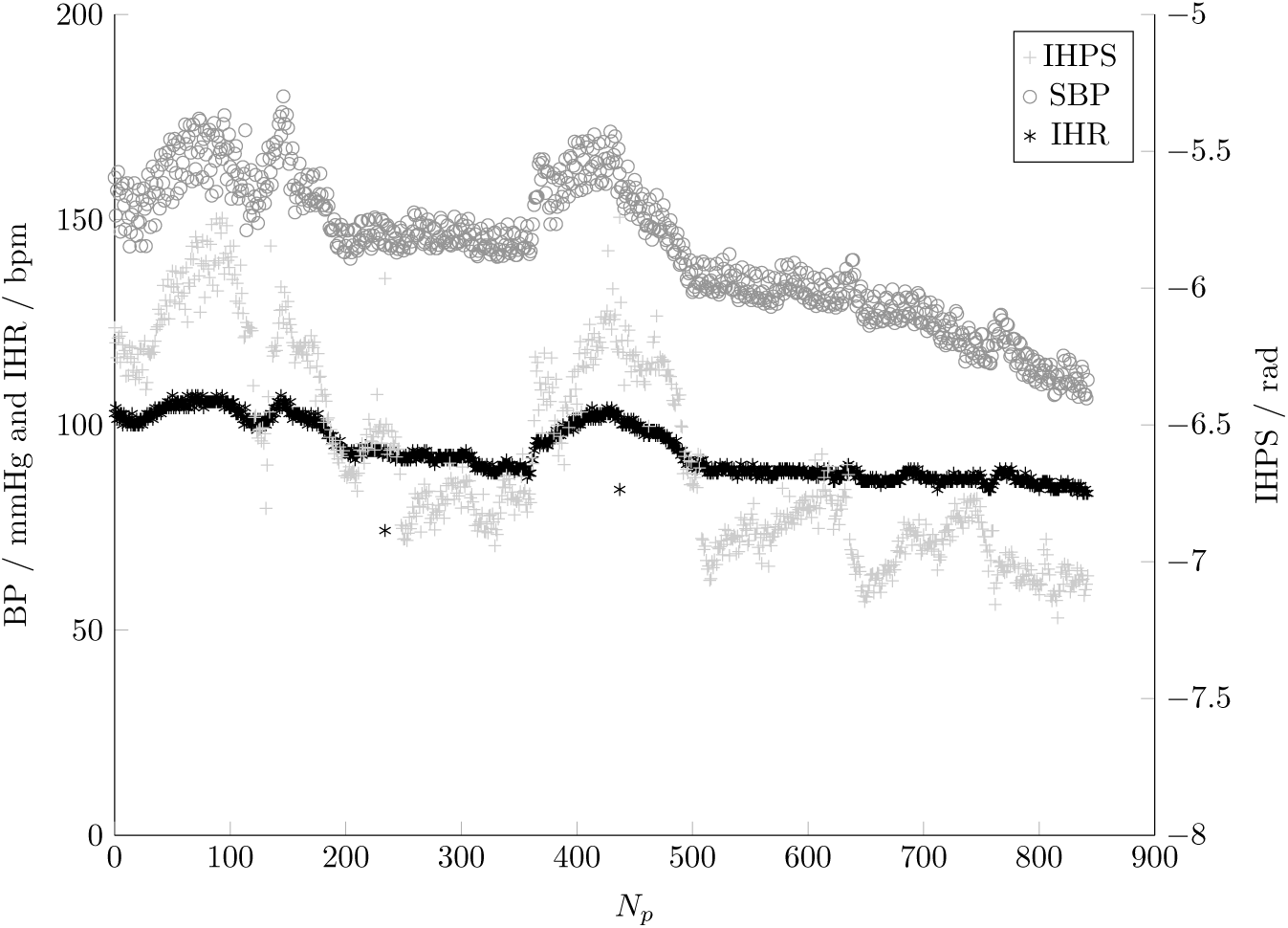
The SBP, the IHPS within the PPG signal and the IHR plotted over *N*_*p*_ = 843 periods. The graph indicates a strong correlation of the signals.

### 2.4. Calculation of the IHPS per Heartbeat

According to [42], the frequency domain analysis of each heartbeat in the PPG signal using the DFT is given by

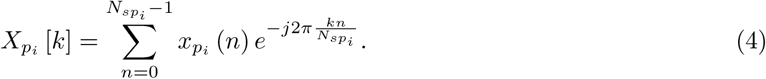

In this equation, 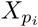 are the complex coefficients resulting from the DFT, 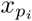(*n*) is the discrete signal being analysed, 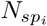 is the number of samples attributed to period *p*_*i*_, *n* is an index to enumerate the samples, *k* is the index of the discrete frequency, and *j* is the imaginary number. The frequency resolution of the DFT is given by 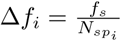, where *f*_*s*_ is the sampling frequency, and 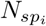 is the number of samples of the *i*^th^ period. The amplitudes (equation 5) and phases (equation 6) can be calculated from the complex numbers 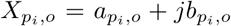, where *o* is the index of the particular harmonic. The amplitude 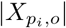 of the *o*^th^ harmonic is obtained by

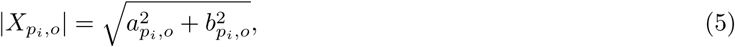

while the phases 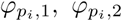 and the relative phase shift 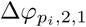 of the fundamental and the first harmonic for a period *p*_*i*_, 1 ≤ *p*_*i*_ ≤ *N*_*p*_ = 843 are computed as:

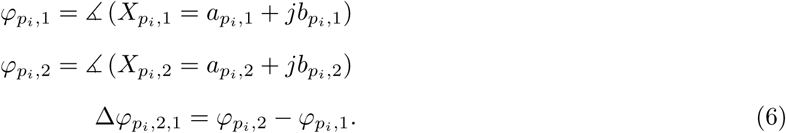

The relative phase-shift is a quantity that describes the difference in phase velocity of the fundamental and the first harmonic at the certain state of arterial dilation. The absolute values of the phase-shifts (and thus also any constant synchronisation delays) play a minor role in the interpretation. Furthermore, we note that the phase-shifts were found to be independent from the IHR that can significantly vary from beat to beat. While this allows for a consistent interpretation of the phase shift throughout the time series, it also paves the way to the application of coherent averaging techniques.

The amplitudes 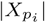 and the phases 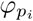 in the typical spectrum of a pulse wave extracted from the PPG signal are shown as double-sided spectrum with the values for the fundamental frequency and the first harmonic highlighted by solid lines in Figure 2. For further calculations, only the phase of the positive frequency spectrum is used. Figure 3 shows the SBP, the IHPS and the IHR that resulted from single beat analysis of the full dataset.

Introducing a quasi-constant frequency resolution allows for a more accurate determination of the harmonics and thus creates better preconditions for correlation. The most prominent options to improve comparability are:

1. Multi-period correlation of IHPS and BP to increase Δ*f*
2. Coherent averaging (CA) without overlap
3. Zero padding and computation of the quasi-continuous spectrum Δ*f*

The main assumption for our method is that any change in the amplitude and offset of the BP is encoded in the frequency domain, where it manifests as phase shift. Altering the time base of the signal in such a way that every single period in a batch of periods has the same period length, as it is done in the case of CA, results to a common fundamental. Since all harmonics are equally stretched, the relative phase-shifts between the fundamental and the harmonics within the periods are preserved.

#### 2.4.1. Multi-Period Correlation of IHPS and BP to Increase Δf

For multi-period analysis, DFT is applied to several successive periods. The filtering done by the multiperiod analysis method smooths the signal in the time domain. By regarding signal sections that span several periods of the PPG signal, the frequency resolution is increased by approximately the number of batched periods, and the relative phase-shift can be determined more precisely compared with single-period analysis. Further, the sizes of the singles batches can by chosen individually and in such a way that the correlation coefficient is maximized. As this analysis can be done with respect to the systole and the diastole, the number of periods being aggregated can largely differ.

#### 2.4.2. Coherent Averaging

Coherent averaging (CA) has been successfully used in similar settings [43, 44]. The idea behind this method is to unify the lengths of a given number of pulse waves in the time domain by stretching the time base of each pulse wave to a common duration, e.g. to the maximal duration found within the whole set. Resampling with a common sample rate will lead to a homogeneous representation that describes every pulse wave with the same number of samples, so that is straightforward to compute the pointwise mean and standard deviation of a batch of pulse waves (see Figure 4). With respect to averaging, the method can easily be varied to evaluate a moving average of pulse wave sequences of some fixed length without any overlapping.

**Figure 4:**
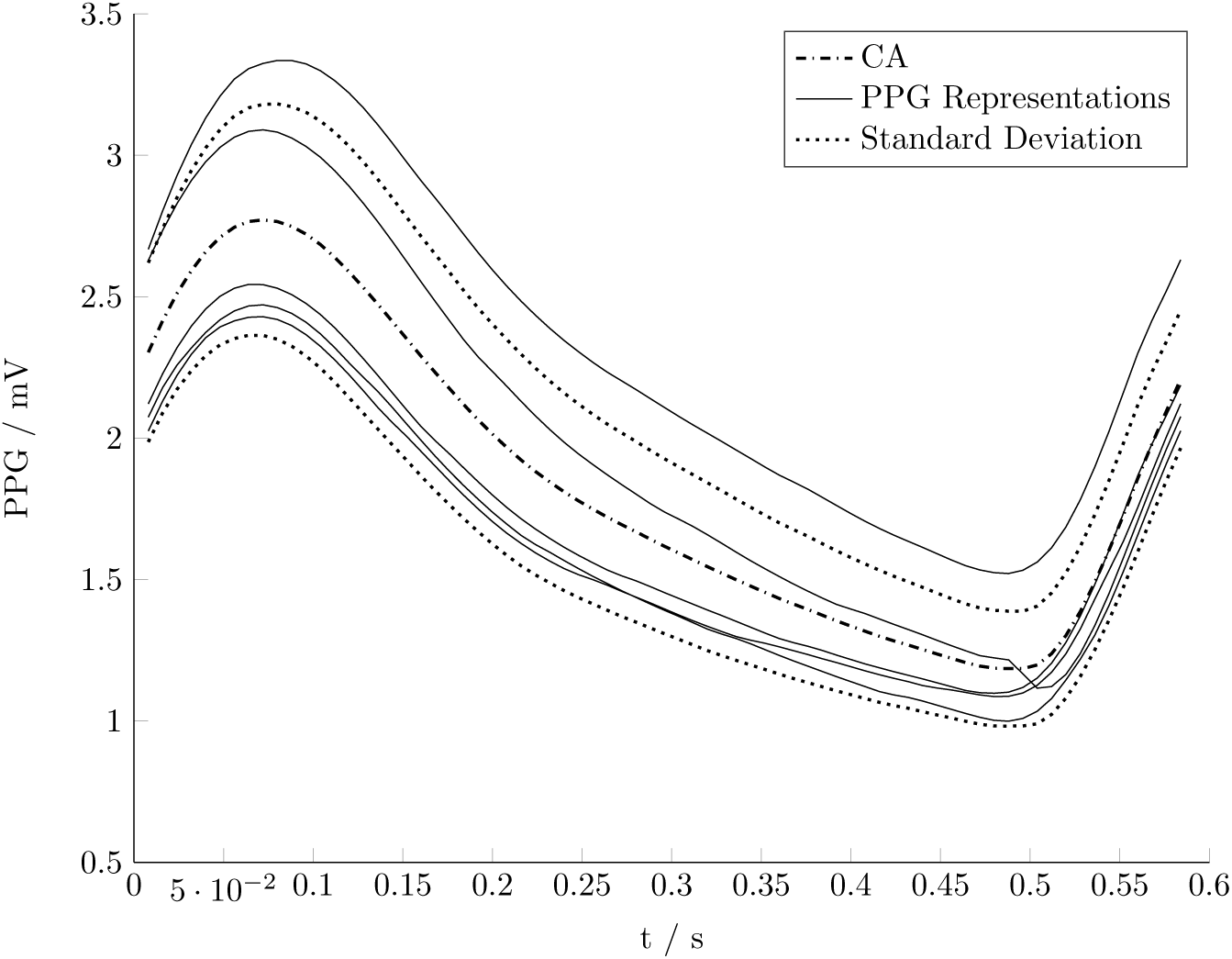
The CA signal calculated as pointwise average from *n* = 5 subsequent pulse waves of the PPG signal stretched to a uniform duration. The pointwise standard deviation can be used to define a band around the averaged signal for outlier treatment.

The advantages of CA lies in a uniform frequency resolution Δ*f*_*CA*_ and an easy outlier treatment.

#### 2.4.3. Zero Padding and Computation of a Quasi-continuous Spectrum

Another common method to improve the frequency resolution is a unification by zero padding each pulse wave to a given length, which also unifies the frequency resolution Δ*f* to a common value. Here, the challenge is to determine the fundamental frequency exactly, since the detection is more difficult due to the leakage effect that comes along with zero padding.

To find the exact harmonics, a quasi-continuous spectrum is computed using interpolation. The resolution of the frequency domain is artificially increased to any desired refinement by computing the quasi-continuous signal

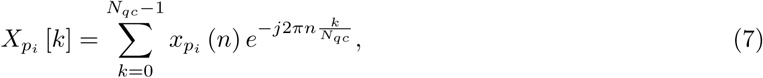

in a refined frequency domain, where 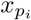(*n*) is the sampled signal for a period *p*_*i*_, *n* is the sample index, *k* is the summation index, and *N*_*qc*_ is the number of values of the quasi-continuous signal. As shown in Figure 5, the fundamental frequency marked with ● and the first harmonic frequency marked with ▪ can be determined via the indices of the associated local amplitude maxima in the quasi-continuous spectrum.

**Figure 5:**
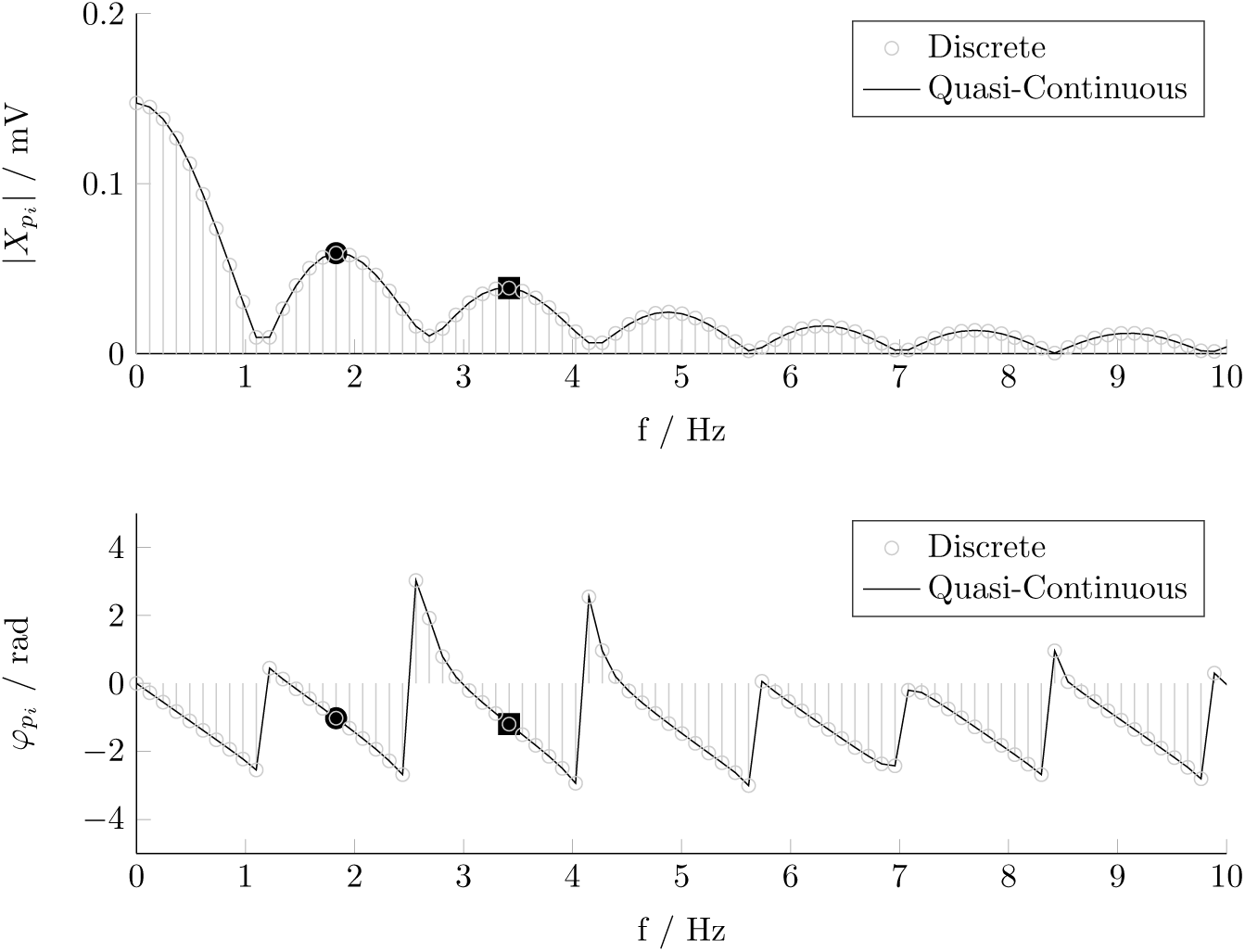
The unilateral amplitude and phase spectra of a PPG signal after unifying the pulse wave lengths by zero padding (with discrete bins) in overlay with the quasi-continuous spectrum (envelope) up to a frequency of 10 Hz. The fundamental frequency marked with ● and the first harmonic marked with ▪ can be determined with high precision by detecting the associated local maxima of the amplitude in the quasi-continuous spectrum.

### 2.5. Determination of the Linear Regression Function between the IHPS and the SBP

To describe the statistical relationship between the SBP and the IHPS it is required to choose a regression function. A scatter plot of the relation, as shown in Figure 6, can help to find out, whether it is promising to conclude a linear regression function

**Figure 6:**
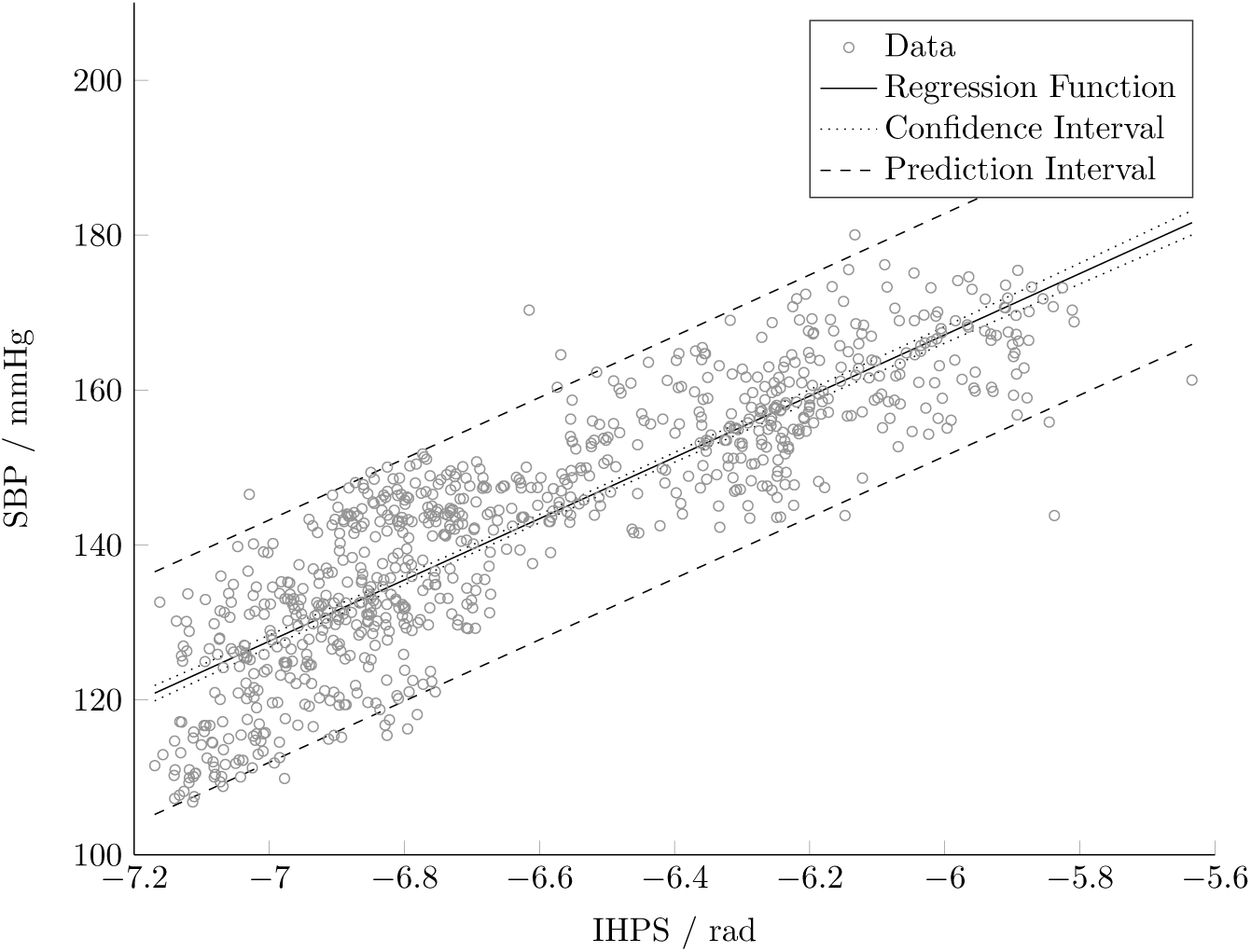
Scatter plot of the relation between the SBP and IHPS obtained by CA with OL with linear regression function, confidence interval and prediction interval determined by linear regression analysis.

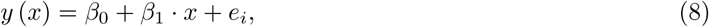

where *β*_0_ is the intercept, *β*_1_ is the slope of the function and *e*_*i*_ the residual of a particular pair with respect to these two parameters. The regression function we used for our regression analysis is given by

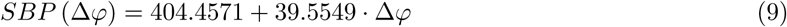

and displayed in Figure 6 together with the confidence interval and the prediction interval for further data with respect to a statistical significance of *α* = 0.05. Note that these results relate to the IHPS obtained by the multi period analysis.

To ensure that the assumptions of linear regression hold for our analysis, it is necessary to check whether the residuals *e*_*i*_ are normally distributed with *e*_*i*_ ∼ 𝒩 ⟨0, *σ*^2^⟩. This holds if (i) the expectation value *ε* ⟨*e*_*i*_⟩ = 0, (ii) the standard deviation 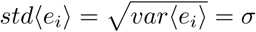 and (iii) the probability plot indicates normality [41]. The requirements (i) and (ii) can be checked by calculating the expectation value and the standard deviation and plotting them together with the residuals against the period index *N*_*p*_, as shown in Figure 7. The probability plot according to requirement (iii) is shown in Figure 8. It indicates normal distribution if the data points spread in a linear fashion around the expectation value and approximate a straight line – and comes out well for our dataset.

**Figure 7:**
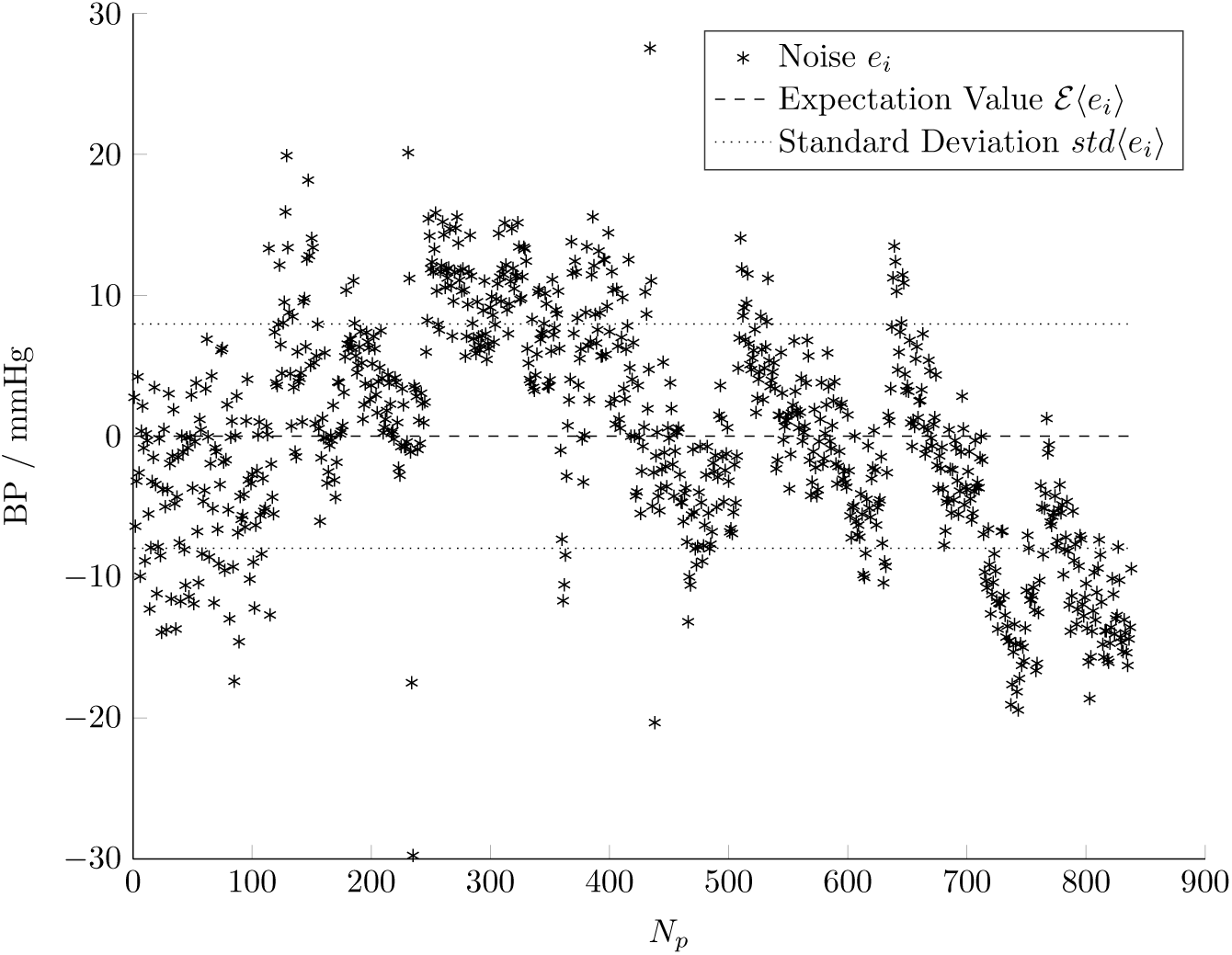
Plot of the residual noise *e*_*i*_ of the regression analysis against the period number with expectation value *ε* ⟨*e*_*i*_⟩ = 0 and standard deviation *std* ⟨*e*_*i*_⟩ = *σ*.

**Figure 8:**
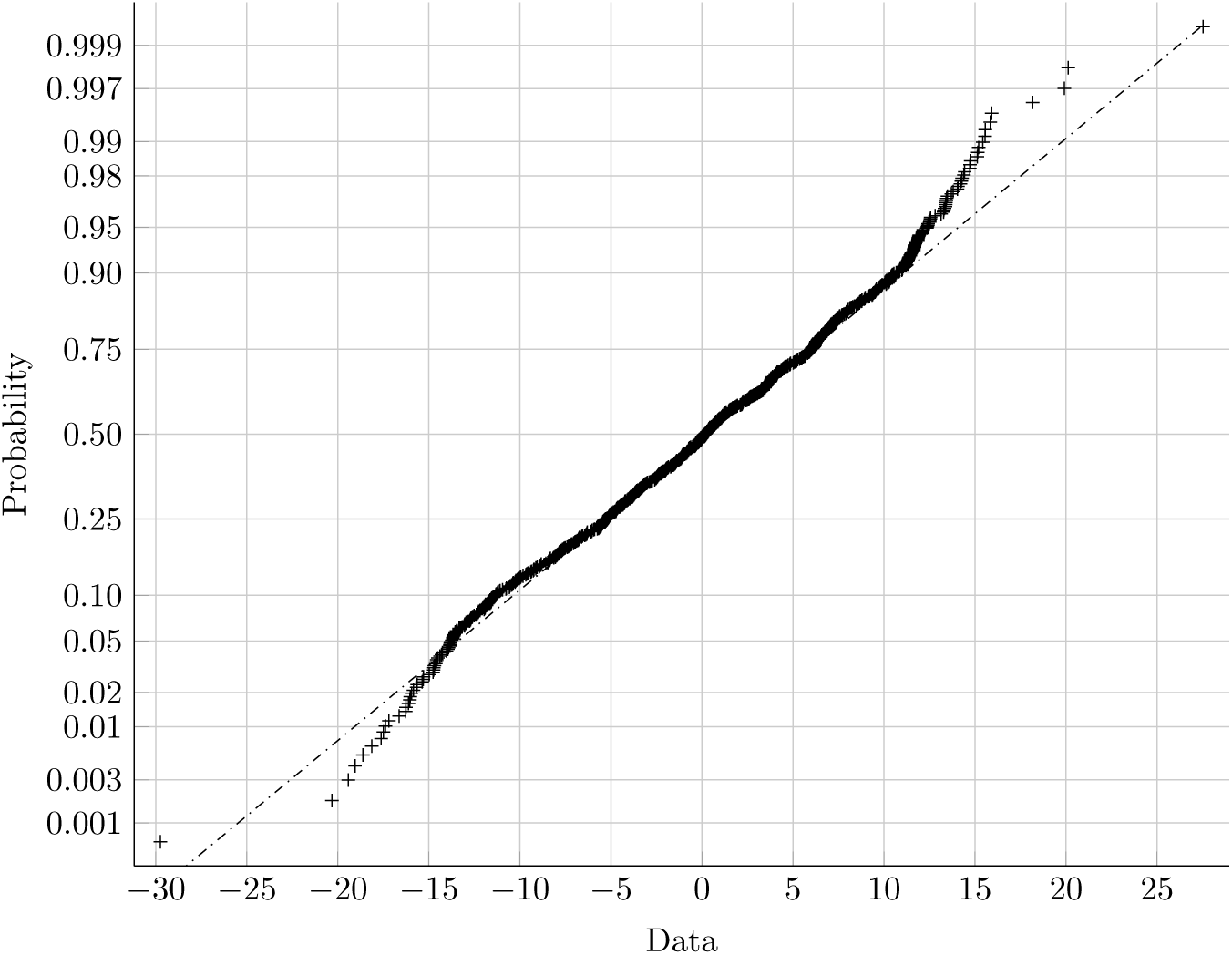
Testing the residuals *e*_*i*_ of our regression analysis for normality to meet the requirements of the regression analysis. Normally distributed data show some dominant linear arrangement and symmetry with respect to the expectation value.

### 2.6. Method Verification by Synthetic PPG Signals from Regression Analysis

The correlation between the BP and the IHPS can in turn also be utilized to simulate the effect of a pressure change on a given PPG signal. Taking some SBP (or DBP) value as argument, the generator function can compute the corresponding modulation of the PPG signal by exploiting the correlation given by the linear regression function (see equations 8 and 9).

Figure 9 shows the results of such a synthetic modulation conducted with a series of systolic pressures given by *sbp* [*k*] = 110*mmHg* + *k ·* 3.5*mmHg*, for {*k* ∈ ℕ | 0 ≤ *k* ≤ 20}. Varying the SBP between 110 and 180 mmHg complied well with the value range found in the previously analysed dataset. The superposed graphs indicate how the PPG signal morphs in response to an increase of the SBP in discrete steps of 3.5 mmHg.

**Figure 9:**
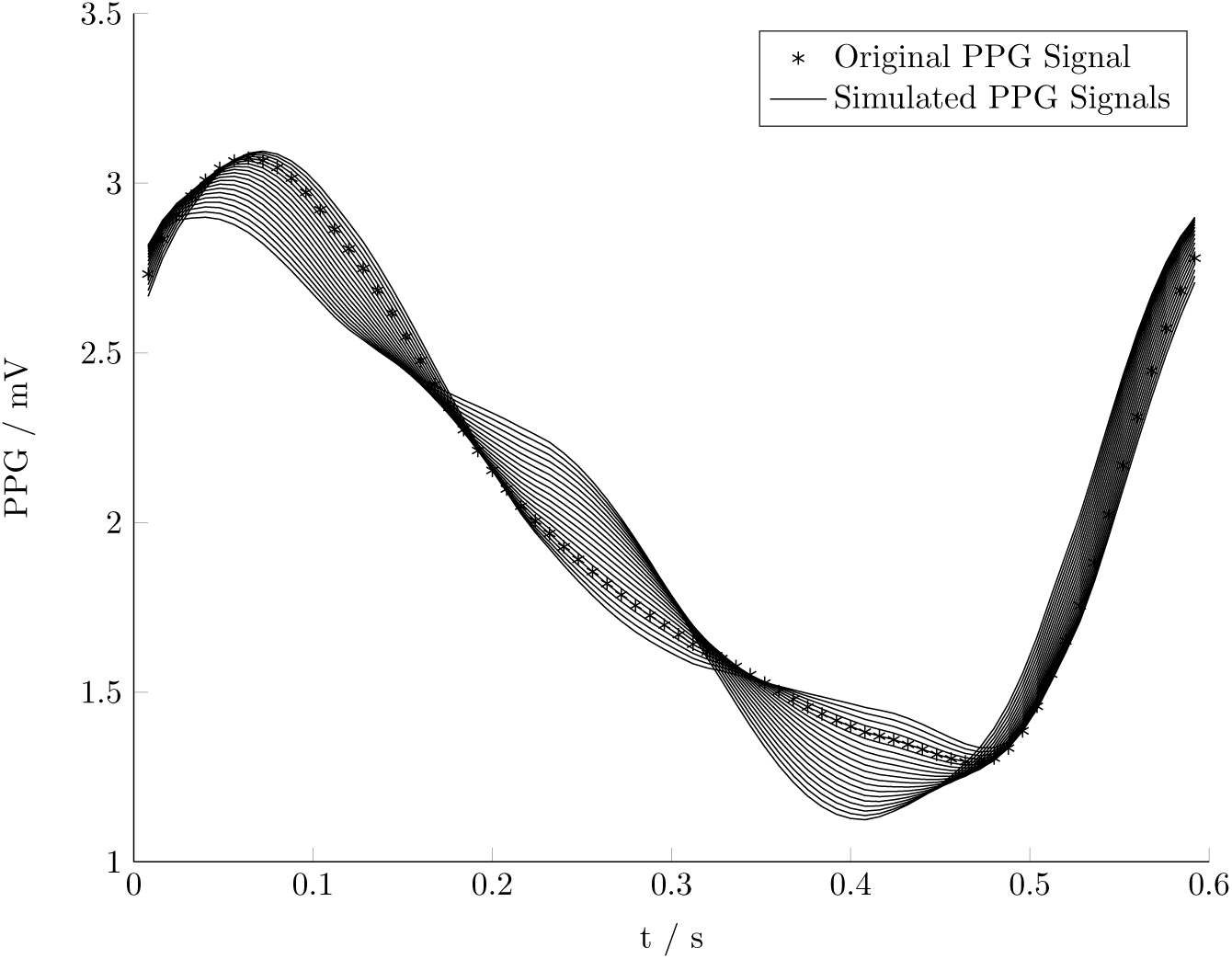
Single period of a measured PPG signal in superposition with 20 simulated PPG signals derived by synthetic modulation of fhe measured PPG signal with a uniform sequence of 20 SBP values ranging between 110 and 180 mmHg for a given regression function.

To validate the phase-shift analysis function of our method, the output of the generator function can be analysed in the same way as a measured PPG signal (with known segmentation). Proper operation is indicated if the phase-shift analysis function exactly finds the IHPS and SBP used for sample generation, provided the parameters of the regression function are known.

## 3. Results

DFT evaluation of a single PPG signal in a beat-to-beat manner with subsequent phase analysis revealed a strong correlation between the IHPS and the IBP. The essential ingredient for the calculation of the IHPS is a precise segmentation of the PPG signal into single pulse waves, which can be accomplished by finding the time indices of the R-peak maxima in the ECG signal, if available. Regarding each pulse wave as full period of a signal, it is straightforward to decompose the signal into a fundamental and its harmonics by DFT. With this pre-processing, specific regression analysis was done by relating the relative phase-shifts between the fundamental and the first harmonic of the single pulse waves to the SBP and the DBP provided by the corresponding IBP signal. Additionally, different post-processing methods were studied that further increased the correlation values. Table 1 shows the obtained results in a concise way and with the strongest correlations highlighted, for comparison the state-of-the-art the correlation coefficients resulting from the PTT method are given side by side. However strongest correlation PTT to BP was observed for the *PTT*_*max*_ which has a mean correlation of approximately 0.63.

Good correlation values with an average of ≈ 0.85 for SBP and DBP over the total signal duration of *N*_*p*_ = 843 periods were determined for single-period analysis without any post-processing. Multi-period analysis in contrast, achieved increased values, and it turned out that the stop criterion used by the procedure to optimize the number of periods combined for multi-period analysis, lead to an average of five periods for the SBP correlation, in contrast to an average of eight periods for the DBP correlation.

The highest correlation values were achieved by regarding batches of subsequent pulse waves, with each pulse wave stretched (and resampled) to the maximum length found in the batch, and then applying CA. For the correlation with SBP the best result (*r* = 0.8945) of all post-processing methods was achieved by CA without overlapping using a batch size of 15 periods, while the correlation with DBP climbed up to *r* = 0.9082 were a batch size of 11 periods was used.

Zero padding with subsequent quasi-continuous spectrum calculation contributed to the lowest correlation values, while demanding the highest calculation effort due to the necessary calculation of the quasi-continuous spectrum.

To support the hypothesis of a linear correlation between IHPS and BP further, the preconditions of linear regression analysis were checked for violation. The analysis of the residuals with respect to the determined regression function unveiled no further functional relation, and the probability plot showed all signs of a normal distribution as well. This confirms that a correct relation was deduced.

Finally, to crosscheck also the algorithm used for phase-shift determination, a synthesised PPG signal gained through modulating a measured PPG pulse wave by means of the previously determined regression function for a wide range of SBP was processed. The analysis reproduced the synthetic input as expected.

## 4. Discussion

In this research, we have proposed and evaluated a novel method for correlating the CBP from harmonic phase-shifts derived from non-invasive peripheral PPG signals. Our comparative study further identified the post-processing method of CA as best method for correlating the IHPS with the SBP, the MBP and also the DBP.

### 4.1. Restrictions of the Investigation

The main limitation for this investigation was the scope of the data regarded. The available dataset had a length of 8m 57s, equivalent to *N*_*p*_ = 843 heartbeats, and a low sample rate of only 125 Hz, contained measurements of only one subject, (iii) originated from an intensive care unit (ICU) patient with no in-depth information available about the state of health, the reason for the stay or any external factors like the conditions of movement or medication. Further limitations of the presented method relate to the algorithms used for analysis: (iv) the phase-shift was only calculated with respect to the fundamental frequency and the first harmonic and (v) an additional ECG signal was used for exact period detection. As the results obtained by the method are very sensitive to the period lengths, period determination had to be done with great care. A segmentation based on the R-peak of the ECG signal proved to be a reliable and robust method.

Despite our very promising results, with regard to the mentioned limitations, the method needs further validation by investigation that is more extensive. Further studies should focus on a refinement of the correlation between the IHPS and the BP. In additon, a linear correlation coefficient might be too restrictive to express a relationship that traces back to arterial stiffness known to have some exponential characteristic with respect to pressure. We are convinced that the presented novel method contains further space for improvement that will increase the correlation coefficient. To harness this potential, it is important to evaluate the phase-shift more quantitatively, under different conditions and with access to suitable measurements that extend over longer time periods and originate from different subjects. Additionally, further research should focus on a method that allows for a reliable period segmentation based on the PPG signal only.

### 4.2. Advantages of the Novel Method

A main advantage of this novel method is the prospect that one standard PPG sensor providing raw data could be sufficient for long time measurement of the BP. In this case the benefit for patients would not only be a simple and reliable measurement method, but also an easy handling that would notably reduce patient load in comparison to the state-of-the-art cuff based or PTT methods, especially for long time measurements. Both, the stress and the restriction of the patients during long-term measurements will be significantly lower than with the standard methods. We expect that by use of this novel method the IBP can be finally estimated from any pulse oximeter providing raw data, which nowadays serves as standard sensor in health monitoring. Furthermore, the analysis of the signal in frequency domain avoids the drawbacks that emerge from time domain analysis, characteristic for the state-of-the-art PTT method. The calculation effort required for the novel method is quite low, so that the algorithms can be easily implemented at the level of common embedded systems. In combination with a small sensor, the entire measurement equipment can be kept very compact.

### 4.3. Uniqueness of the Solutions

The analysis of only one PPG signal for the determination of the BP in the frequency domain is an essential feature of the novel method and thereby significantly improves the measuring conditions by the simple measurement setup using only one sensor at the fingertip. Furthermore, the analysis of the signal in frequency domain is not dependent on the afore said drawbacks presented in the state-of-the-art PTT method emerging in time domain analysis, which is also due to the fact that the determination of the phase-shift is a relative quantity in comparison to the absolute time measurement of the PTT method. Beside the simplicity of the measurement setup and the analysis algorithms, the correlation of the single signal analysis is as high as the state-of-the-art PTT correlation values. According to its simplicity, we assume that the novel method will improve the measurement setup and interpretation of BP data of hypertensive patients in near feature significantly. Analysing other datasets with the phase-shift algorithm have further supported this method and provide similar results. In comparison to the results obtained by means of PTT in the correlation between either PTT and the SBP/DBP, the method presented here is at least as good as most other results [5, 24–27, 45].

### 4.4. Outlook to Further Investigations

The quasi-continuous analysis of the PPG signal in frequency domain and its good results in correlation with the BP shows the potential of the novel method for non-invasive CBP determination by means of a PPG signal in the frequency domain. By further improving the method through the use of better analytical methods and other transformations or relations, it should be possible to obtain continuous results using this novel method.

## Data Availability

All data is available free on UCI Machine Learning Repository of the University of California, Irvine, which is part of the “PhysioNet” data base

https://archive.ics.uci.edu/ml/index.php

## Acknowledgement

This work was funded by the State of Baden Wuerttemberg, Germany, Ministry of Science, Research and Arts within the scope of Cooperative Research Training Group.

## Conflict of interest/funding/ethical approval

1. Conflicts of Interest: None
2. Funding: State of Baden Wuerttemberg, Germany, Ministry of Science, Research and Arts
3. Ethical Approval: Not required

